# Drug Repurposing of potential drug targets for treatment of COVID-19

**DOI:** 10.1101/2021.06.02.21258223

**Authors:** Rajaneesh K. Gupta, Enyinna L. Nwachuku, Benjamin E. Zusman, Ruchira M. Jha, Ava M. Puccio

## Abstract

Drug repurposing has the potential to bring existing de-risked drugs for effective intervention in an ongoing pandemic—COVID-19 that has infected over 131 million, with 2.8 million people succumbing to the illness globally (as of April 04, 2021). We have used a novel `gene signature’-based drug repositioning strategy by applying widely accepted gene ranking algorithms to prioritize the FDA approved or under trial drugs. We mined publically available RNA sequencing (RNA-Seq) data using CLC Genomics Workbench 20 (QIAGEN) and identified 283 differentially expressed genes (FDR<0.05, log2FC>1) after a meta-analysis of three independent studies which were based on severe acute respiratory syndrome-related coronavirus 2 (SARS-CoV-2) infection in primary human airway epithelial cells. Ingenuity Pathway Analysis (IPA) revealed that SARS-CoV-2 activated key canonical pathways and gene networks that intricately regulate general anti-viral as well as specific inflammatory pathways. Drug database, extracted from the Metacore and IPA, identified 15 drug targets (with information on COVID-19 pathogenesis) with 46 existing drugs as potential-novel candidates for repurposing for COVID-19 treatment. We found 35 novel drugs that inhibit targets (ALPL, CXCL8, and IL6) already in clinical trials for COVID-19. Also, we found 6 existing drugs against 4 potential anti-COVID-19 targets (CCL20, CSF3, CXCL1, CXCL10) that might have novel anti-COVID-19 indications. Finally, these drug targets were computationally prioritized based on gene ranking algorithms, which revealed CXCL10 as the common and strongest candidate with 2 existing drugs. Furthermore, the list of 283 SARS-CoV-2-associated proteins could be valuable not only as anti-COVID-19 targets but also useful for COVID-19 biomarker development.

## Introduction

A novel coronavirus (CoV) began at the end of 2019 in Wuhan, China: it infected over seventy thousand individuals within the first fifty days of the epidemic with 1800 individuals succumbing to the disease [1]. The World Health Organization (WHO) declared a public health emergency of international concern on January 30 that escalated to a pandemic on March 11, 2020 [2]. Amino acid and nucleotide sequencing studies confirmed this new CoV belonged to the same species as severe acute respiratory syndrome coronavirus-1 (SARS-CoV-1) [3]. The International Committee on Taxonomy of Viruses (ICTV) named this new CoV as severe acute respiratory syndrome coronavirus 2 (SARS-CoV-2) and the disease as CoV disease 2019 (COVID-19) [4]. SARS-CoV-2 consists of four structural proteins including spike (S) glycoprotein, envelope (E) protein, membrane (M) protein, and nucleocapsid (N) protein. Host cell binding and entry are mediated by the S protein. The S1 subunit of the S protein can mediate entry into human respiratory epithelial cells by interacting with cell surface receptors like angiotensin-converting enzyme 2 (ACE2). Entry also requires S protein priming by cellular proteases like Type II transmembrane serine protease (TMPRSS2), resulting in S protein cleavage and fusion of viral and airway cellular membranes [5]. Patients with infected airway epithelium can present with a range of symptoms including fever, cough, and shortness of breath, often culminating in acute lung injury, acute respiratory distress syndrome, pulmonary failure, and even death [6]. As of April 04, 2021, more than 30 million cases of COVID-19 have been reported with over 1.6% mortality rate in the U.S. alone [7]. According to WHO’s International Clinical Trials Registry Platform (ICTRP), 8,936 projects including 5,126 interventional studies are registered related to COVID-19 across the globe (searched on April 04, 2021) [8]. With the possible exception of dexamethasone and anti-inflammatory therapy, the currently available medications have questionable and limited efficacy against improving outcomes after COVID-19. The WHO SOLIDARITY trial (an international clinical trial to seek an effective treatment for COVID-19) results have shown that remdesivir (FDA approved drug for COVID-19 treatment) has little or no efficacy on mortality rate in hospitalized patients with COVID-19 [9]. Therefore, there is an urgent need to further identify novel medical therapies both for preventive and therapeutic use. Drug repurposing could significantly curtail the time and cost of development compared to *de novo* drug discovery, as toxicity and safety data are often available from former clinical trial phases [10]. *In silico* approaches based on functional annotations proffer novel testable hypotheses for systematic drug repurposing [11].

RNA-sequencing (RNA-Seq) based transcriptional profiling of SARS-CoV-2 infected airway epithelial cells provides a remarkable opportunity for understanding the relationship between infection-triggered gene expression signature and viral pathogenesis. A meta-analysis using differentially expressed genes (DEGs) obtained from RNA-Seq studies has the potential to advance our understanding of SARS-CoV-2 pathogenesis and facilitates the process of anti-COVID-19 drug repurposing. This current study systematically and quantitatively combined analysis of multiple RNA-Seq studies using a meta-analysis approach. This helps decrease the inconsistency in individual studies by increasing the sample size and statistical power to enlist more robust SARS-CoV-2-associated genes [12]. Furthermore, existing, approved drugs with opposing effects on these SARS-CoV-2-associated genes could have the potential to reverse COVID-19 symptoms. Therefore, we employed an *in silico* approach to discover potential anti-COVID-19 drug targets and suggest a priority for repurposing drugs which are FDA approved or under clinical trial investigation as potential COVID-19 therapies. Finally, this work provides insights into the untested but potential gene targets in relation to COVID-19 pathogenesis.

## Materials and methods

### Study search

This study utilized the CLC Genomics Workbench 20.0.3 database [13] to search for deposited RNA-Sequencing experiments related to SARS-CoV-2 infections that matched the following criteria: only whole transcriptome studies; experiments carried out in human airway epithelial cells infected with SARS-CoV-2 *in vitro*; and availability of the raw data (.fastq files) for each sample. We searched on Dec 03, 2020 with search terms: “SARS-CoV-2”, “SARS-CoV-2 pandemic”, “SARS-CoV-2 host response”, “SARS-CoV-2 transcriptome”, “COVID-19”, “COVID-19 pandemic”, “COVID-19 host response”, and “COVID-19 transcriptome”. We identified three independent studies shown in Table 1 and selected the reads associated with SARS-CoV-2 or mock-treated airway epithelial cells. Details of selected reads and experimental conditions are given in <u>Table S1</u>.

**Table 1.** Detailed information regarding the three selected studies associated with SARS-CoV-2.

| Study title | Dataset | Platform | Sample size | Hours post infection | Airway epithelium |
| --- | --- | --- | --- | --- | --- |
| Infection of human nasal epithelial cells with SARS-CoV-2 and a 382-nt deletion isolate lacking ORF8 reveals similar viral kinetics and host transcriptional profiles | GSE162131 | Illumina HiSeq 4000 | SARS-CoV-2 infected: 6 | 24 | Human nasal epithelium |
|  |  |  | SARS-CoV-2 infected: 6 | 72 |  |
|  |  |  | Mock infected: 6 | N/A |  |
| Primary human airway epithelial cultures infected with SARS-CoV-2 | GSE153970 | Illumina NovaSeq 6000 | SARS-CoV-2 infected: 6 | 48 | Primary human airway epithelium |
|  |  |  | Mock infected: 6 | N/A |  |
| Transcriptional response to SARS-CoV-2 infection | GSE147507 | NextSeq 500 | SARS-CoV-2 infected: 12 | 24 | Primary human bronchial epithelium |
|  |  |  | Mock infected: 12 | N/A |  |

### RNA-sequencing data collection, processing, and analysis

Raw reads (.fastq files) were downloaded from CLC genomics workbench 20.0.3 [13]. A quality check for raw reads was performed. Low-quality bases (Phred score<20) and adapters were excluded. Trimmed reads were mapped to the human_sequence_hg38 reference genome to verify valid reads. Reads were extracted, counted, and normalized. Genes with false discovery rate (FDR)-adjusted *p*-values<0.05 and log2 fold change (log2FC)>1 were (Figure 1).

The gene datasets were analyzed for disease and disorders, molecular and cellular functions, and canonical pathways using Ingenuity Pathway Analysis (IPA) version 60467501 [14]. Only experimentally observed studies associated to the human species were used for IPA analysis. Fisher’s exact test was used to calculate a *p*-value determining the probability that overlap with each canonical pathway is due to chance alone. In contrast to canonical pathways, which are relatively immutable in IPA, gene networks were also generated *de novo* in IPA based on the list of genes that are imported. IPA takes Network Eligible molecules from the gene list, searches the Ingenuity Knowledge Base, and uses a network algorithm to draw connections between molecules based on biological function [14].

**Figure 1.**
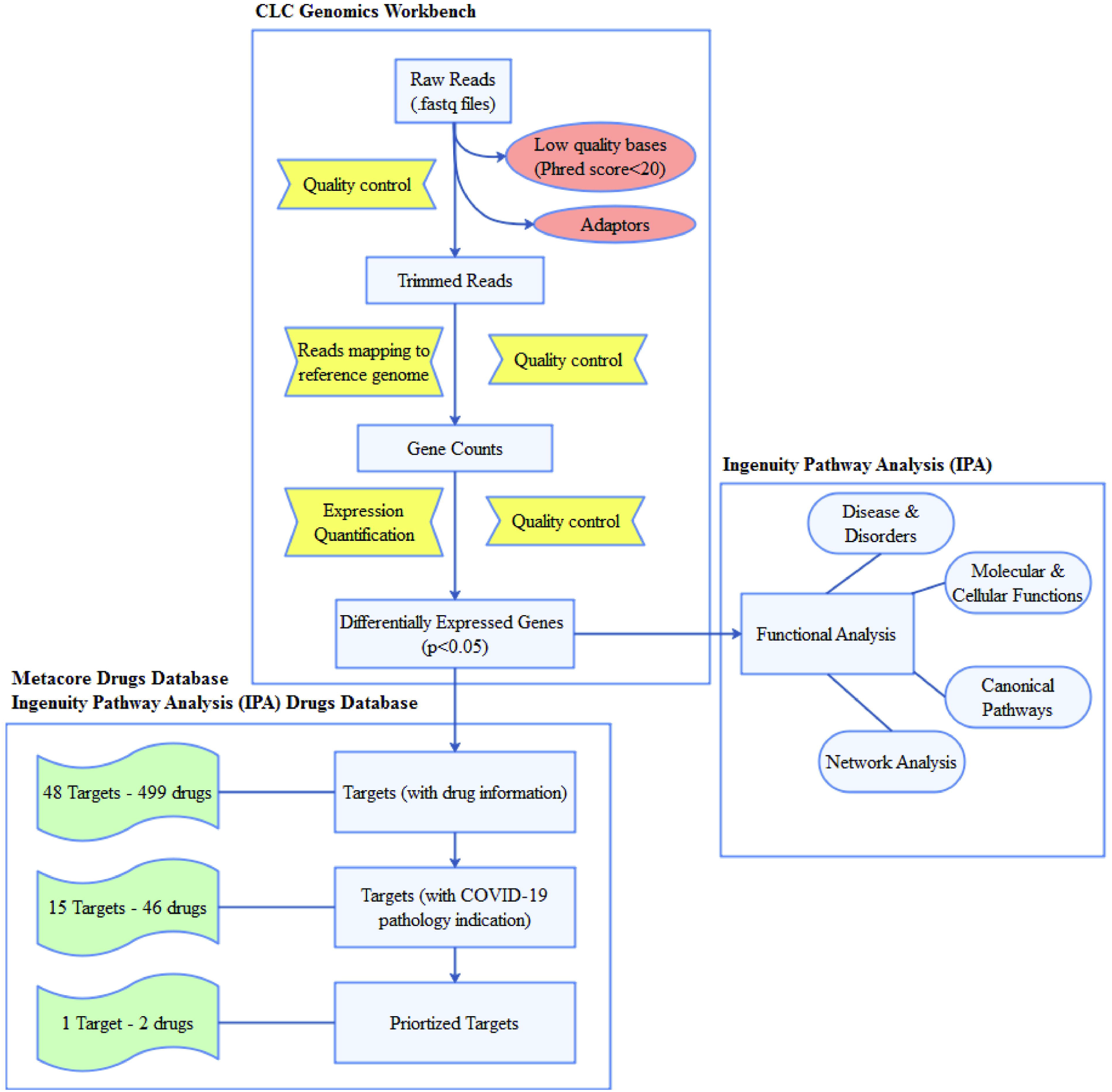
Flow-chart of the drug repositioning strategy for COVID-19 based on SARS-CoV-2 triggered gene expression signature in airway epithelium.

### Mapping SARS-CoV-2-related DEGs to FDA approved/ clinical drugs

To establish a link between SARS-CoV-2-related genes to drugs, we used a commercial drug database of Metacore version 20.3 build 70200 from Clarivate Analytics [15] and IPA [14]. To highlight the most promising drugs that might be repurposed for COVID-19 treatment, only drug targets that were either FDA approved or had been examined in clinical trials were selected. From these drug databases, we extracted information on drug name, drug target, and drugs’ mechanism of action.

### Information on pathogenesis and the drugs’ modes of action for anti-SARS-CoV-2 drug repurposing

The list of all genes involved in the pathogenesis of COVID-19 was extracted (as of April 04, 2021) from the IPA database. Information on specific target activity and the effect of each target on disease/function were examined. Target pathogenesis information with the drugs’ mechanisms of action retrieved from the drug databases was used to rationally shortlist promising anti-SARS-CoV-2 drugs.

### Computational analysis of candidate drug targets and repurposed drugs

Validation of the anti-SARS-CoV-2 drug targets derived from the SARS-CoV-2 infection induced transcriptome of human airway epithelium, was performed using two globally-accepted disease gene prioritizing tools [16]. Gene prioritization tools utilize mathematical and computational models of disease to filter the original set of genes based on functional similarity (Toppgene tool, https://toppgene.cchmc.org) and, topological features in protein-protein interaction (Toppnet tool, https://toppgene.cchmc.org) to the training genes. The Online Mendelian Inheritance in Man (OMIM) database (http://www.omim.org) was searched for genes (training genes) whose inhibition or activation significantly affects the progression of SARS-CoV pathogenesis in human patients.

## Results

### SARS-CoV-2 induced transcriptome in human airway epithelium

This meta-analysis identified 283 DEGs at FDR<0.05 and log2FC>1 (<u>Table S2</u>). IPA identified 27 disease and disorders (p-value<0.05) (Figure 2A; <u>Table S3A</u>), 21 cellular and molecular functions (p-value<0.05) (Figure 2B; <u>Table S3B</u>), and 275 canonical pathways (p-value<0.05) (Fig 2C; <u>Table S3C</u>). Infectious disease and Cell movement, cell death/survival, and cell signaling were the major disease and cellular functions respectively compromised following SARS-CoV-2 infection in respiratory epithelium. Canonical pathway analysis showed the role of IL17A in psoriasis, the role of IL17F in inflammation in airways, and chronic obstructive pulmonary disease (COPD) signaling pathways were severely affected by SARS-CoV-2 infection. Overall, this suggests the infectious inflammatory nature of host response in respiratory epithelial cells that could culminate in cell death and other respiratory problems in response to SARS-CoV-2 infection, consistent with what is seen in clinical practice.

**Figure 2.**
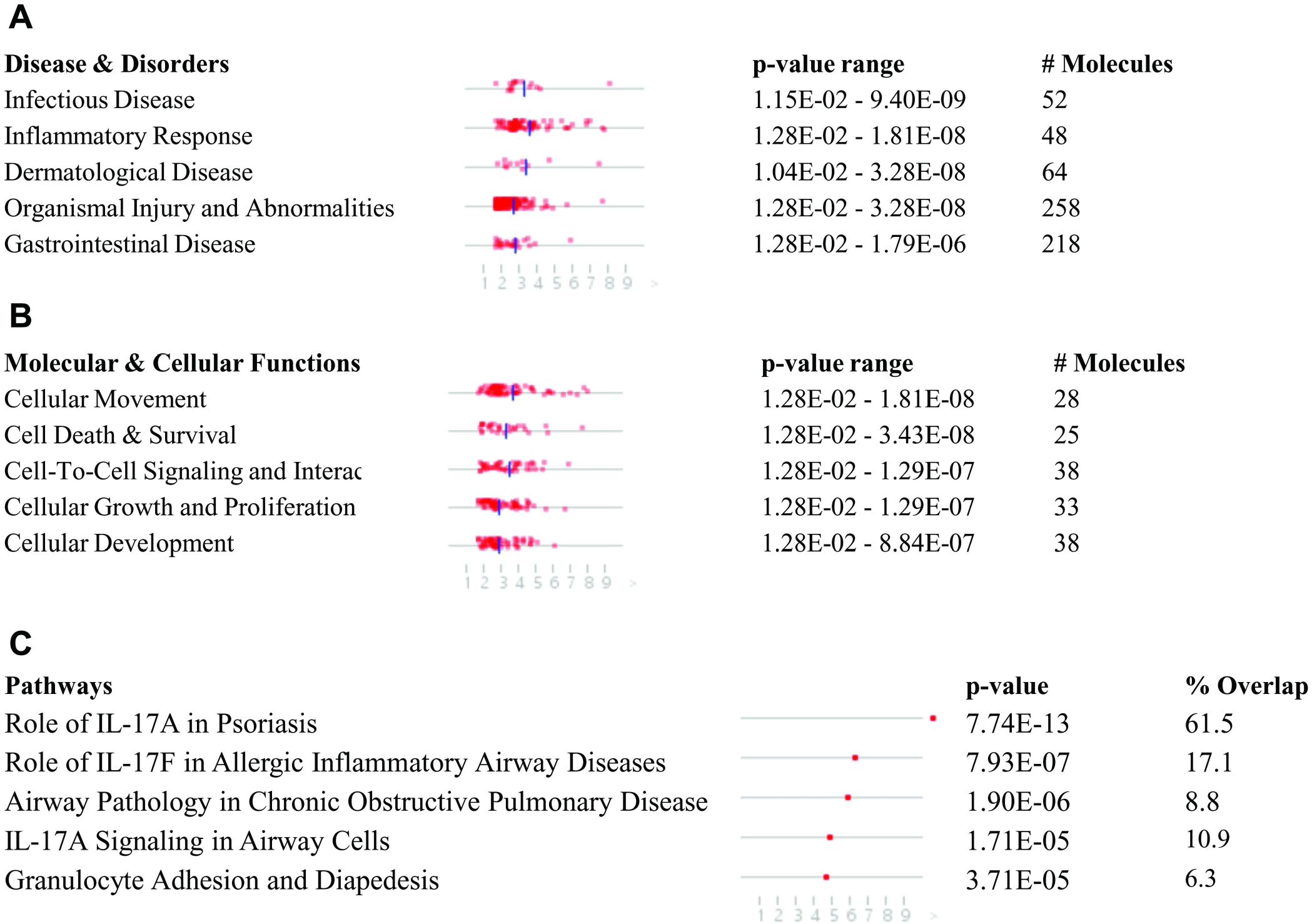
List of top 5 (A) Disease and disorders, (B) cellular and molecular functions, and (C) canonical pathways that were enriched in the meta-analysis of three RNA-sequencing assays associated with SARS-CoV-2 infected (*in vitro*) airway epithelium.

The IPA network algorithm created 25 connection network between molecules based on biological function (<u>Table S3D</u>) and ranked them as per the network score. The score takes into account the number of Network Eligible molecules in the network and its size, as well as the total number of Network Eligible molecules analyzed and the total number of molecules in the Ingenuity Knowledge Base that could potentially be included in networks. The higher the score, the lower the probability of finding the observed number of Network Eligible molecules in a given network by random chance. Figure 3 shows three most relevant networks. Network-1 (score=35; Figure 3A) was associated with 23 genes of our dataset and was associated with ‘Cellular movement, hematological system development and function, immune cell trafficking’. This network was characterized by key cytokines IL6, IL16, CXCL1, CXCL3, CXCL5, CXCL6, CXCL8, CXCL10, and CCL20. Network-2 (score=25; Figure 3B) was associated with ‘Cellular development, cellular growth and proliferation, embryonic development’ with transcription factors (POU6F1, MAFB), enzymes (RSAD2, AKR1C1, GBP5, TRIM36, GSTA2), and cytokines (CD70, CSF2, CSF3) as hub molecules. Network-3 (score=19; Figure 3C) was associated with ‘Cell death and survival, cell morphology, organismal development’. This network was centralized on transcription factors (SOX6, ZNF114), enzymes (CMBL, CSGALNACT1, MAN1C1), and others (BCL2A1, CDRT1, SYNE1).

**Figure 3.**
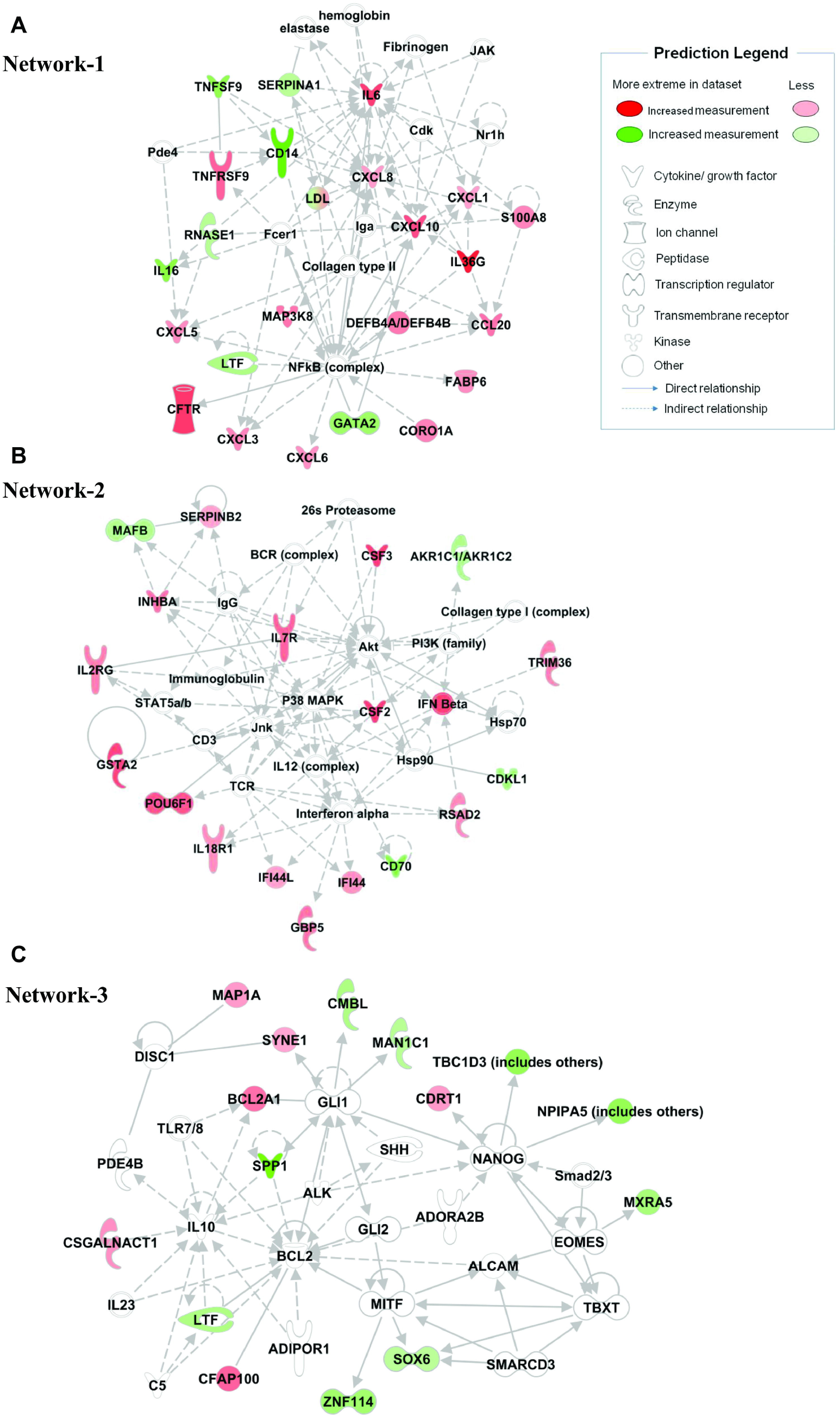
Generated molecular network based on differential expression of genes of SARS-CoV-2 infected (in vitro) airway epithelium associated with (A) ‘Cellular movement, hematological system development and function, immune cell trafficking’, (B) ‘Cellular development, cellular growth and proliferation, embryonic development’, and (C) ‘Cell death and survival, cell morphology, organismal development’ using ingenuity knowledge database. Coloring is based on the expression values of the genes, down-regulation in green and, up-regulation in red. Genes with no coloring are added from ingenuity knowledge database. Direct and indirect relationships are shown by solid and dashed lines, respectively. The arrow indicates specific directionality of interactions.

### Existing drugs with possible anti-COVID-19 indication derived from knowledge of drugs’ modes of action and COVID-19 pathogenesis

The Metacore and IPA databases were searched using ENSEMBL IDs for the abovementioned 283 protein targets. Fifteen DEGs (with information on COVID-19 pathogenesis) were related to 46 FDA approved or clinical trial drugs (with data on drugs’ modes of action) (<u>Table S4</u>), supporting their potential candidate nature against SARS-CoV-2. Three of these 15 targets - ALPL (drug: zinc sulfate), CXCL8 (drug: BMS-986253), IL6 (drug: clazakizumab, siltuximab, tocilizumab) are currently under clinical trial for COVID-19. This supports the ability of our approach to identify anti-SARS-CoV-2 drugs and suggests its potential to discover novel anti-COVID-19 indications for existing drugs. We also identified 35 drugs targeting ALPL, CXCL8, and IL6 that were not previously tested against COVID-19 symptoms and could be repurposed for anti-SARS-CoV-2 management. We additionally found 4 potential targets (CCL20, CSF3, CXCL1, CXCL10) with 6 existing drugs that could be repurposed for the treatment of COVID-19. Eight further targets (CXCL6, IFI44, IFI44L, RSAD2, S100A8, SPRR2A, SYNE1, XAF1) associated with COVID-19 pathogenesis were identified, but drugs of any therapies that are either approved for another indication or being studied in a clinical trial.

### Computational ranking of drug targets based on functional similarity and topological features in protein-protein interaction to SARS-CoV pathology genes

We subsequently implemented publicly available online gene ranking algorithms to prioritize the anti-SARS-CoV-2 targets to find out which of the drugs targeting 15 genes are the most reliable. An extensive search through OMIM literature was performed to identify reliable training genes for the SARS-CoVs infection pathology. We found 4 genes (training genes) that are directly associated with SARS-CoVs pathology (ACE2; Gene MIM number 300335, AGTR1; Gene MIM number 106165, DPP4; Gene MIM number 102720, TMPRSS2; Gene MIM number 602060). TopGene has generated a similarity score for each annotation of each test gene by comparing to the enriched terms in the training set of genes. The final prioritized gene list is then computed based on the aggregated values of all similarity scores (Table 2, <u>Table S5</u>). Another online gene ranking algorithm, ToppNet has mapped training and test set genes to protein-protein interaction network. Scoring and ranking of test set genes based on relative location to all of the training set genes using global network-distance measures in the protein-protein interaction network (PPIN) (Table 2, <u>Table S5</u>). CXCL10 was the topmost prioritized target with the maximum score analyzed by ToppGene and ToppNet. CXCL10 is involved in the regulation of inflammatory and immune responses (Figure 4) and may be a potential candidate target for treating COVID-19 related lung pathology.

**Table 2.** SARS-CoV-2 triggered gene expression signature revealed potential anti-COVID-19 drug targets from existing approved and clinical trial drugs.

| ENSEMBL ID | Target symbol | PMID/NCT No. | Target score |  | No. of drugs |
| --- | --- | --- | --- | --- | --- |
|  |  |  | ToppGene | ToppNet |  |
| ENSG00000169245 | CXCL10 | 32427582, 32612617, 32446778, 32407669 | 4.70E-01 | 1.44E-03 | 2 |
| ENSG00000169429 | CXCL8 | NCT04347226, 32427582, 32612617, 32471782, 32407669 | 4.61E-01 | 1.03E-04 | 16 |
| ENSG00000136244 | IL6 | 32475230, 32501293, 32427582, 32612617, 32612615, 32428990, 32471782, 32475810, 32446778, 32439209, 32344321, | 4.09E-01 | 8.90E-06 | 18 |
|  |  | NCT04345445, NCT04423042,<br>NCT04403685, NCT04356937,<br>NCT04403685, NCT04412772 |  |  |  |
| ENSG00000108342 | CSF3 | 32612617 | 3.60E-01 | 6.49E-06 | 1 |
| ENSG00000163739 | CXCL1 | 32407669 | 3.39E-01 | 2.12E-05 | 1 |
| ENSG00000115009 | CCL20 | 32407669 | 3.35E-01 | 2.74E-05 | 2 |
| ENSG00000162551 | ALPL | NCT04370782 | 2.60E-01 | 1.42E-05 | 6 |

**Figure 4.**
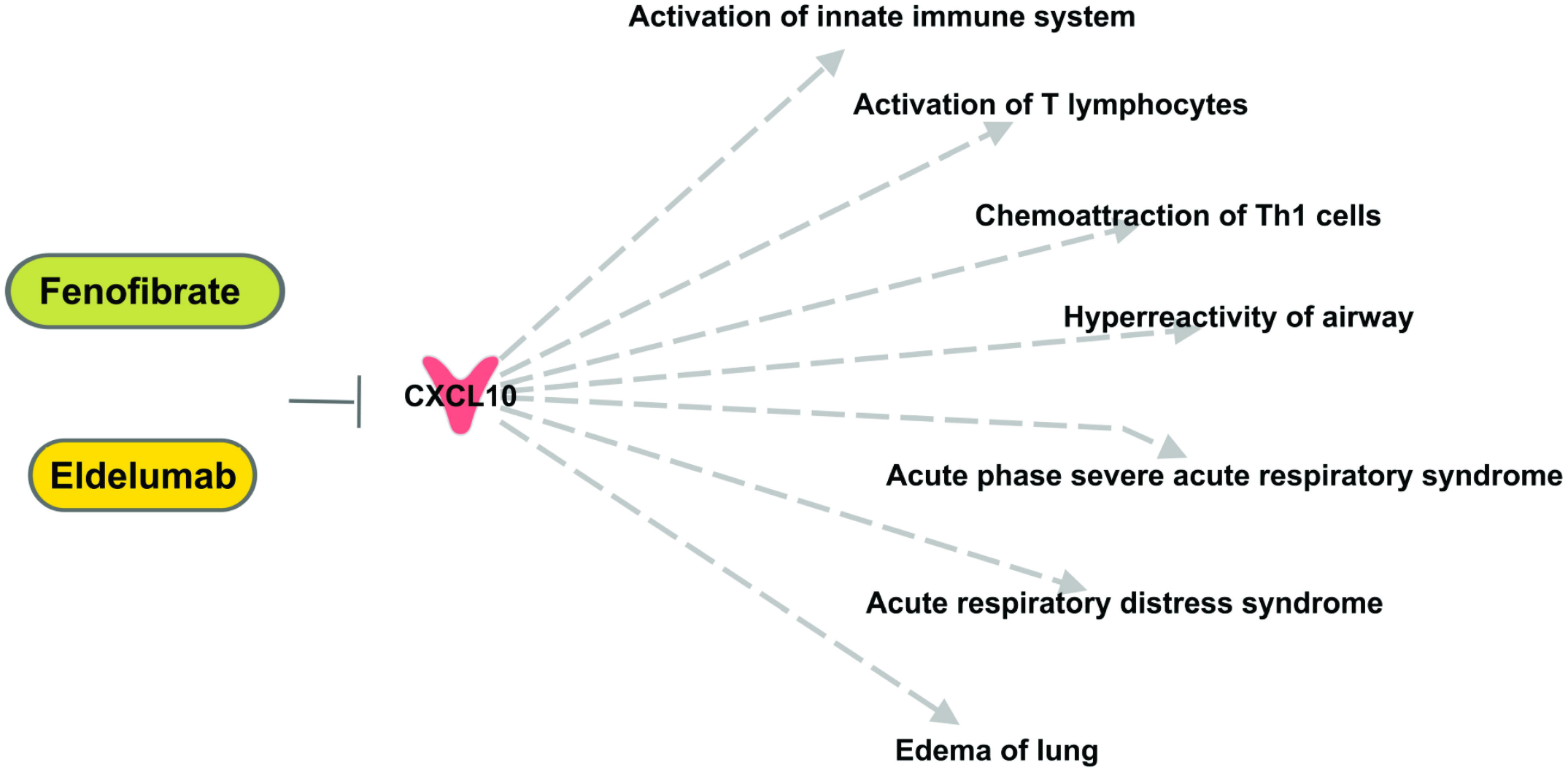
Top anti-SARS-CoV-2 target, CXCL10, affect inflammatory and immune responses. Eldelumab and Fenofibrate may be assessed for their effects in modulating COVID-19-related lung pathology.

## Discussion

Despite the growing understanding of the mechanisms of SARS-CoV-2 pathology, interventions for treatment of COVID-19 are limited. Most importantly, the identity of critical targets in infected cells that can be modulated in the early/middle stage of disease are not well known. Such interventions could blunt the development of more severe symptoms, hospitalization and death. In the current study, we performed a quantitative meta-analysis of three independent RNA-Seq studies to identify SARS-CoV-2-triggered genes in the transcriptome of human respiratory epithelial cells. Data mining and subsequent extension of bioinformatics analysis of RNA-Seq data revealed 283 DEGs, 15 of which are the target of 48 existing drugs, which could be repurposed for the treatment of COVID-19. Three of these 15 targets IL6, ALPL, and CXCL8 are currently being studied as therapies for hospitalized COVID-19 patients (Table 2). Our drug discovery pipeline suggested an additional 35 existing drugs (<u>Table S4</u>) targeting these molecules that can be repurposed. Importantly, our meta-analysis identified 4 potential novel targets (CCL20, CSF3, CXCL1, CXCL10) associated with COVID-19 pathogenesis, targeted by 6 existing drugs that have not been studied in clinical trials for treatment of COVID-19 and could be repurposed. An additional 8 targets (CXCL6, IFI44, IFI44L, RSAD2, S100A8, SPRR2A, SYNE1, XAF1) are associated with COVID-19 pathogenesis, but do not have therapies targeting them available for potential repurposing. The list of 283 SARS-CoV-2-related proteins could be useful not only as potential anti-COVID-19 targets, but could also be considered COVID-19 biomarkers of disease progression and/or response to therapy.

The interaction network among these 283 gene points to core hub genes that could be responsible for altered molecular and cellular functions ‘cell movement’, ‘cell death & survival’ and ‘cell signaling/proliferation’ associated with enriched diseases ‘infection’ and ‘inflammatory response’ following SARS-CoV-2 infection in airway epithelium. Inflammatory response in viral infection is a double-edged sword: Although inflammation is necessary to orchestrate the recruitment and coordination of immune cells at the infection site, over-stimulation of the inflammatory response results in a surge in cytokines and could culminate into secondary injury or destruction of the respiratory epithelium. Our IPA analysis showed that activation of cytokines IL6, IL16, CXCL1, CXCL3, CXCL5, CXCL6, CXCL8, CXCL10, and CCL20 are a hallmark signature of cytokine response in COVID-19 patients [17,18]. We also found increased levels of key pro-apoptotic factors like BCL2A1, LTF, SOX6, SPP1, and SYNE1. Li et al. has recently detected apoptosis in bronchial and lung epithelial cells during the initial exudative phase (day 2-4 post infection) of SARS-CoV-2 infection in humanized ACE2 transgenic mice [19]. These pro-apoptotic factors could represent key nodes associated with epithelial cell death and subsequent diffuse alveolar damage (a critical feature of acute lung injury). Further, canonical pathway analysis suggested an enriched ‘IL17 signaling’ pathway. IL17 family cytokine levels are associated with respiratory viral infections [20]. It regulates key inflammatory cytokines like CSF3, TNFα, IL6, IL-1β, CXCL10, IL-8, MIP2A, MMPs that control granulopoiesis and recruitment of neutrophils, fever, chemoattraction, and tissue damage and remodeling [21]. There is a growing body of evidence supporting the role of IL17 in COVID-19 pathogenesis [20,22]. However, further investigation is needed to characterize the role of IL17 blockade in the management of COVID-19. Several existing drugs targeting IL17 could be tested for COVID-19 therapies (e.g. secukinumab [approved for arthritis, psoriasis, spondylitis], ABY-035 [for psoriasis; NCT02690142], BCD-085 [for arthritis; NCT03598751, for spondylitis; NCT03447704; for psoriasis; NCT03390101], bimekizumab [for arthritis; NCT04009499], CJM112 [for hidradenitis suppurativa; NCT02421172]).

We further used globally accepted ranking algorithms to prioritize the drug targets. These algorithms analyzed the targets’ functional and topological similarity to established COVID-19 pathogenesis genes and both scored CXCL10 the highest priority drug target. CXCL10 has been implicated in SARS-CoV [23] and other viral infections such as rhinovirus, respiratory syncytial virus (RSV), Coxsackie virus, hepatitis virus B and C, Ebola, dengue (DENV), and equine infectious anemia virus (EIAV) [24]. Ichikawa et al. have shown that mice deficient in CXCL10 or its receptor CXCR3 have decreased lung injury severity and increased survival after viral and non-viral lung injury [25]. CXCL10 levels were positively correlated with the extent of organ damage and pathogen burden [26]. Interestingly, increased levels of CXCL10 were found in plasma samples of patients with COVID-19 who died [17]. Furthermore, Chua et al. have highlighted the up-regulation of chemokines including CXCL10 in airway epithelial cells extracted from patients with moderate or critical COVID-19 [27]. Of note, a clinical trial assessing the use of serial CXCL10 measurement as a clinical decision support for patients with COVID-19 recently completed enrollment (NCT04389645) supporting the notion that this may be an actionable target in this patient population.

One possible CXCL10 targeting therapy is Eldelumab. Eldelumab (MDX-1100 or BMS-936557) is a fully humanized antibody (type IgG1 kappa developed by Bristol-Myers Squibb and Medarex) targeting CXCL10 [28] that is being studied as a therapy for multiple autoimmune and autoinflammatory diseases including rheumatoid arthritis (NCT01017367), ulcerative colitis (NCT00656890, NCT01294410), and Crohn’s disease (NCT01466374). It is hypothesized that by binding to CXCL10, eldelumab blocks immune cell migration into the epithelium and modulates the impact of CXCL10 on epithelial cell survival [29]. The safety profile seems favorable, with the most commonly reported treatment-related adverse event being a mild to moderate headache (5% patients) with 10 or 20 mg/kg (intravenous infusion) doses [30], though the optimal treatment dose and safety profile would need to be investigated specifically in the COVID-19 patient population.

Fenofibrate is another therapy that may reduce CXCL10 activity and is hypothesized to have therapeutic activity in inflammatory diseases like Crohn’s disease [31]. Fenofibrate may also decrease the expression of other cytokines (IL17, CCL2, and CCL20) implicated in COVID-19 pathology [31]. As a peroxisome proliferator-activated receptor alpha (PPARα) agonist, fenofibrate may also prevents phospholipid accumulation within SARS-CoV-2 infected cells, blocking viral replication [32]. It may also suppress microvascular inflammation and apoptosis through inhibition of nuclear factor-κB and activation of adenosine monophosphate (AMP)-activated protein kinase [33], suggesting the fenofibrate may further have favorable systemic anti-inflammatory and endothelial effects.

In conclusion, systematic analyses of the SARS-CoV-2 triggered gene signature in airway epithelium revealed 15 protein targets linked to 46 existing drugs. This include 35 drugs modifying the activity of molecules already being studied as therapeutic targets for COVID-19 disease (IL6, ALPL, CXCL8 targets) and which could likely be repurposed for a similar aim. Our study also found 4 additional targets (CCL20, CSF3, CXCL1, CXCL10) with existing therapies that have yet to be trialed in the COVID-19 patient population. CXCL10 appears to be a particular strong candidate based on high target scores and the availability of two existing drugs inhibiting the action of this cytokine. Our study has several limitations. This investigation is based on *in vitro* RNA-Seq data, resulting in an under-appreciation of significant inter-cellular signaling that may occur differently in the human body. Furthermore, our computational approach is limited as a tool for evaluating drugs to be repurposed because most available computational tools are used for small molecule drugs only. However, given the pressing need for effective targeted therapies for the treatment of COVID-19, further studies are crucially needed to experimentally validate these results and, if promising, rapidly transition to clinical trials.

## Data Availability

All data are available as supplementary.

## Acknowledgement

Authors thank to Molecular Biology Information Service of the Health Sciences Library System, University of Pittsburgh for support on data analysis by CLC Workbench Genomics 20 and Ingenuity pathway analysis (IPA).

## Supporting information

**Table S1. Details of selected reads and experimental conditions for each GEO dataset**

**Table S2. A complete list of differential expressed genes in airway epithelium after SARS-CoV-2 infection identified in meta-analysis**.

**Table S3. List of (A) Disease and disorders, (B) cellular and molecular functions, (C) canonical pathways, and gene networks that were enriched in the meta-analysis of three RNA-sequencing assays associated with SARS-CoV-2 infected (*in vitro*) airway epithelium**

**Table S4. Potential anti-SARS-CoV-2 drug targets with existing FDA approved or clinical trial drugs**.

**Table S5. Ranking of the potential AD drug targets using (A) ToppGene and (B) ToppNet web tools**.

